# Predicting 30-Day In-Hospital Mortality in Surgical Patients: A Logistic Regression Model Using Comprehensive Perioperative Data

**DOI:** 10.1101/2024.05.18.24307573

**Authors:** Jonathan Hofmann, Andrew Bouras, Dhruv Patel, Nitin Chetla, Nia Balaji, Michael Boulis

## Abstract

**Background:** Accurate prediction of postoperative outcomes, particularly 30-day in-hospital mortality, is crucial for improving surgical planning, patient counseling, and resource allocation. This study aimed to develop and validate a logistic regression model to predict 30-day in-hospital mortality using comprehensive perioperative data from the INSPIRE dataset.

**Methods:** We conducted a retrospective analysis of the INSPIRE dataset, comprising approximately 130,000 surgical cases from Seoul National University Hospital between 2011 and 2020. The primary objective was to develop a logistic regression model using preoperative and intraoperative variables. Key predictors included demographic information, clinical variables, laboratory values, and the emergency status of the operation. Missing data were addressed through multiple imputation, and feature selection was performed using univariate analysis and clinical judgment. The model was validated using cross-validation and assessed for performance using ROC AUC and precision-recall AUC metrics.

**Results:** The logistic regression model demonstrated high predictive accuracy, with an ROC AUC of 0.978 and a precision-recall AUC of 0.958. Significant predictors of 30-day in-hospital mortality included emergency status of the operation (OR: 1.56), preoperative prothrombin time (PT/INR) (OR: 1.53), potassium levels (OR: 1.49), body mass index (BMI) (OR: 1.37), serum sodium (OR: 1.11), creatinine levels (OR: 1.04), and albumin levels (OR: 0.85).

**Conclusion:** This study successfully developed and validated a logistic regression model to predict 30-day in-hospital mortality using comprehensive perioperative data. The model’s high predictive accuracy and reliance on routinely collected clinical and laboratory data enhance its feasibility for integration into existing clinical workflows, providing real-time risk assessments to healthcare providers. Future research should focus on external validation in diverse clinical settings and prospective studies to assess the practical impact of this predictive model.

## 1 Introduction

Predicting postoperative outcomes is a critical component of perioperative medicine, enabling health-care providers to identify high-risk patients and tailor perioperative management strategies accordingly.

In particular, accurately forecasting 30-day in-hospital mortality can significantly impact surgical planning, patient counseling, and resource allocation (Khuri et al., 1997). Despite advancements in surgical techniques and perioperative care, mortality within 30 days post-surgery remains a pertinent concern, necessitating robust predictive models that can incorporate a variety of patient-specific factors (Gibbs et al., 1999).

The utilization of machine learning (ML) techniques in healthcare has garnered considerable interest, offering the potential to enhance predictive accuracy by leveraging large datasets and complex variable interactions (Deist et al., 2018). However, the interpretability and clinical applicability of these models remain paramount. Logistic regression, a well-established statistical method, provides a balance between predictive performance and interpretability, making it a suitable choice for clinical decision support (Chiew et al., 2019).

This study aims to develop and validate a logistic regression model to predict 30-day in-hospital mortality using a comprehensive dataset of perioperative information from the INSPIRE dataset. The dataset encompasses approximately 130,000 surgical cases from Seoul National University Hospital (SNUH) between 2011 and 2020, providing a rich source of preoperative and intraoperative variables (Lim et al., 2023; Goldberger et al., 2000).

Key predictors in our model include demographic information, clinical variables, laboratory values, and the emergency status of the operation. Special attention is given to preoperative albumin levels due to their strong correlation with patient nutritional status and overall health (Caironi et al., 2009). By incorporating these variables, the model seeks to offer reliable risk assessments that can inform clinical decisions and improve patient outcomes.

Through rigorous methodological approaches, including multiple imputation for missing data, cross-validation for model stability, and feature selection using both statistical and clinical criteria, we aim to develop a robust predictive tool. The findings of this study will contribute to the growing body of literature on perioperative risk prediction, providing valuable insights for enhancing surgical care and patient management.

## 2 Literature Review

The prediction of postoperative outcomes, particularly 30-day in-hospital mortality, has been an area of active research in perioperative medicine. Various approaches, ranging from traditional risk assessment models to advanced machine learning techniques, have been explored to enhance the accuracy and applicability of predictive tools. This literature review synthesizes key findings from existing studies, highlighting the evolution of predictive models and the integration of diverse clinical variables.

### 2.1 Traditional Risk Assessment Models

Traditional risk assessment models, such as the American Society of Anesthesiologists (ASA) physical status classification, have long been used to stratify surgical risk. The ASA classification, although widely adopted, has demonstrated limited predictive accuracy, with reported receiver operating characteristic (ROC) area under the curve (AUC) values typically ranging from 0.60 to 0.75 (Khuri et al., 1997). Other traditional models, such as the Surgical Apgar Score and the POSSUM (Physiological and Operative Severity Score for the enumeration of Mortality and morbidity), also face similar limitations in predictive power and often lack granularity in incorporating comprehensive perioperative variables (Merkow et al., 2013).

### 2.2 Preoperative Variables and Outcomes

Preoperative variables play a crucial role in predicting postoperative outcomes. Studies have consistently identified factors such as age, sex, comorbidities, and nutritional status as significant predictors of surgical risk. Preoperative albumin levels, a marker of nutritional status, have been highlighted for their strong association with postoperative morbidity and mortality (Gibbs et al., 1999). Caironi et al (2009) discussed the clinical implications of albumin in intensive care, underscoring its value as a predictor of outcomes in surgical patients (Caironi et al., 2009).

### 2.3 Machine Learning Approaches

The advent of machine learning has opened new avenues for developing predictive models in health-care. Machine learning algorithms, including random forests, gradient boosting machines, and neural networks, have demonstrated improved predictive accuracy over traditional methods. For example, Chiew et al. (2019) utilized machine learning methods to predict postsurgical mortality and ICU admission, reporting ROC AUC values around 0.85 to 0.90, significantly higher than traditional models (Chiew et al., 2019). Similarly, Deist et al. (2018) compared various machine learning classifiers for outcome prediction in (chemo)radiotherapy, highlighting the empirical superiority of these techniques (Deist et al., 2018).

### 2.4 Logistic Regression in Predictive Modeling

Despite the rise of complex machine learning models, logistic regression remains a widely used and valuable method in clinical research due to its interpretability and ease of implementation. Logistic regression models have been effectively employed to predict various clinical outcomes, including mortality and complications in surgical patients. Hill et al. (2018) utilized logistic regression to predict in-hospital mortality using electronic medical record data, demonstrating that logistic regression, when properly implemented, can yield high predictive accuracy (Hill et al., 2018).

### 2.5 Feature Selection and Model Validation

Feature selection and model validation are critical components in developing robust predictive models. Techniques such as Lasso regularization help prevent overfitting by penalizing the absolute size of regression coefficients, thus simplifying the model without sacrificing predictive power. Multiple imputation methods, such as the chained equations approach, are used to handle missing data, ensuring the completeness and reliability of the dataset. Cross-validation, a method to assess the model’s stability and generalizability, is frequently employed to ensure robust performance across different subsets of the data (Tseng et al., 2020).

### 2.6 Comparative Studies

Comparative studies of traditional risk models and machine learning approaches underscore the benefits and limitations of each method. While machine learning models often exhibit superior predictive performance, logistic regression models offer greater interpretability and clinical relevance. For instance, studies comparing the ASA classification with machine learning techniques have consistently shown higher predictive accuracy with machine learning, yet logistic regression models remain preferred for their simplicity and ease of integration into clinical practice (Chiew et al., 2019); (Hill et al., 2018).

## 3 Methods

This study is a retrospective analysis of perioperative data from the INSPIRE dataset, which includes approximately 130,000 surgical cases that underwent anesthesia at a major academic institution in South Korea between 2011 and 2020 (Lim et al., 2023; Goldberger et al., 2000). The primary objective was to develop and validate a machine learning model capable of predicting 30-day in-hospital mortality using a combination of preoperative and intraoperative variables. Data for this research were sourced from the INSPIRE dataset, comprising anonymized records of patients who underwent various surgical procedures under different types of anesthesia at Seoul National University Hospital (SNUH). The dataset was curated from the clinical data warehouse of SNUH, focusing on patients administered general, neuraxial, regional, or monitored anesthesia care. Exclusion criteria included patients younger than 18 or older than 90 at the time of operation and those who refused or were unable to consent to data disclosure. This dataset provides a comprehensive range of perioperative information, including patient demographics, details of the surgical and anesthesia procedures, laboratory results, and postoperative outcomes.

The analysis concentrated on variables closely linked with the risk of 30-day in-hospital mortality. A significant emphasis was placed on preoperative albumin levels due to their notable correlation with patient nutritional status and overall health. Additional key predictors examined included demographic variables such as age and sex; clinical variables including body mass index (BMI), the emergency status of the operation (emop), and the American Society of Anesthesiologists (ASA) physical status; and a range of laboratory values—hemoglobin, platelet count, white blood cell count, partial thromboplastin time (PTT), prothrombin time (PT/INR), glucose, blood urea nitrogen (BUN), creatinine, sodium, potassium, and liver function tests (AST, ALT). Moreover, the ICD-10 coding for procedures (icd10 cm) was integrated into the analysis to adjust for the type and complexity of the surgical procedures, recognizing that more complex surgeries could pose higher risks and influence patient outcomes differently.

To justify the sample size, we conducted a power analysis using the G*Power software. Given the anticipated effect size, an alpha level of 0.05, and a power of 0.80, we determined that a minimum sample size of 10,000 patients would be necessary to detect significant predictors of 30-day in-hospital mortality with sufficient power. The large sample size of approximately 130,000 cases in our dataset exceeds this requirement, ensuring the robustness of our statistical analyses and the reliability of the model’s predictions. Missing data were addressed through multiple imputation using the chained equations method (MICE). This approach allows for the imputation of missing values based on the relationships between observed data points, ensuring that the imputed values are consistent with the overall data distribution. We performed sensitivity analyses to evaluate the impact of imputed data on our results, comparing models with and without imputed values to confirm the stability of our findings.

We employed logistic regression to predict the probability of 30-day in-hospital mortality. The dataset was divided into a training set (70%) and a validation set (30%) to train and subsequently validate the model, ensuring robustness and generalizability of the findings. The following diagnostics and validation procedures were applied: we assessed multicollinearity using the Variance Inflation Factor (VIF). Variables with a VIF greater than 10 were considered highly collinear and were either transformed or excluded from the model. We performed the Hosmer-Lemeshow test to evaluate the goodness-of-fit of the logistic regression model, ensuring that the predicted probabilities align well with the observed outcomes. Diagnostic plots of residuals (e.g., standardized residuals, leverage plots) were examined to identify any influential data points or patterns indicating model misspecification. The model’s performance was evaluated using the area under the receiver operating characteristic (ROC) curve (AUC) and precision-recall curve metrics. These measures provide insights into the sensitivity and specificity of the model across various threshold settings, highlighting its accuracy in distinguishing between patients who survived and those who did not within 30 days post-operation. We performed five-fold cross-validation to assess the model’s stability and robustness.

Feature selection was conducted through a combination of univariate analysis and expert clinical judgment. Each potential predictor variable was individually tested for its association with 30-day in-hospital mortality using logistic regression. Variables with p-values less than 0.05 were considered for inclusion in the multivariate model. Variables identified as significant in the univariate analysis were further assessed for their clinical relevance and potential impact on surgical outcomes based on existing literature and expert opinion. Significant variables from the univariate analysis were entered into a multivariate logistic regression model. Stepwise selection methods (both forward and backward) were used to identify the most parsimonious model, ensuring that only variables with independent predictive value were retained. To prevent overfitting and enhance model generalizability, we applied Lasso (Least Absolute Shrinkage and Selection Operator) regularization, which penalizes the absolute size of the regression coefficients, thereby shrinking less important variables’ coefficients towards zero and effectively performing variable selection.

By following these rigorous methodological steps, we aimed to develop a robust, reliable model that accurately predicts 30-day in-hospital mortality based on preoperative factors, ultimately enhancing preoperative assessments and improving patient management strategies in surgical settings.

For access to the code used in this analysis, please refer to the following repository: https://github.com/hofmannj0n/biomedical-research.

## 4 Results

The results of the logistic regression model demonstrated substantial predictive accuracy for 30-day inhospital mortality. The model’s coefficients were interpreted as odds ratios (OR) with 95% confidence intervals (CI), providing a clearer understanding of the relationship between each predictor and the outcome. The emergency status of the operation was the most influential predictor, with an OR of 1.56 (95% CI: 1.44-1.68, p <0.001), highlighting the increased risk associated with emergency surgeries. PT/INR had an OR of 1.53 (95% CI: 1.48-1.58, p <0.001), indicating a 53% increase in mortality odds per unit increase. Preoperative potassium levels had an OR of 1.49 (95% CI: 1.44-1.54, p <0.001), suggesting a 49% increase in mortality odds per unit increase. BMI was another significant predictor, with an OR of 1.37 (95% CI: 1.33-1.41, p <0.001). Preoperative serum sodium had an OR of 1.11 (95% CI: 1.09-1.13, p <0.001), indicating that each unit increase in sodium level was associated with an 11% increase in the odds of mortality. Preoperative creatinine had an OR of 1.04 (95% CI: 1.03-1.05, p <0.001), suggesting a 4% increase in mortality odds per unit increase in creatinine level. Preoperative albumin was a significant protective factor with an OR of 0.85 (95% CI: 0.84-0.87, p <0.001), implying a 15% decrease in the odds of mortality per unit increase in albumin level.

The logistic regression model’s performance metrics were robust, with an area under the receiver operating characteristic (ROC) curve (AUC) of 0.978 and a precision-recall AUC of 0.958. These high values indicate excellent discrimination between patients who survived and those who did not within the 30-day postoperative period. Cross-validation results further confirmed the model’s reliability, with an average ROC AUC of 0.987 and an average precision-recall AUC of 0.975 across five folds. The data was divided into five subsets for cross-validation, with each subset used once as a test set while the remaining four subsets were used for training, ensuring that all data points were used for both training and validation.

For a comprehensive evaluation, we compared our model’s performance with other predictive models in the literature. Traditional models, such as the ASA physical status classification, have demonstrated lower predictive accuracy with ROC AUC values typically ranging from 0.60 to 0.75. Recent studies utilizing machine learning techniques, such as random forests and gradient boosting machines, have reported ROC AUC values around 0.85 to 0.90 (Chiew et al., 2019, Deist et al., 2018, Hill et al., 2018). Our logistic regression model outperformed these models, showcasing superior predictive power.

The feature importance analysis revealed that the emergency status of the operation (OR: 1.56, 95% CI: 1.44-1.68, p <0.001) was the most influential predictor of mortality, followed by PT/INR (OR: 1.53, 95% CI: 1.48-1.58, p <0.001), preoperative potassium levels (OR: 1.49, 95% CI: 1.44-1.54, p <0.001), BMI (OR: 1.37, 95% CI: 1.33-1.41, p <0.001), preoperative serum sodium (OR: 1.11, 95% CI: 1.09-1.13, p <0.001), creatinine levels (OR: 1.04, 95% CI: 1.03-1.05, p <0.001), and albumin (OR: 0.85, 95% CI: 0.84-0.87, p <0.001).

**Table 1:** Ranked Feature Importance from Logistic Regression Model.

| Feature | Odds Ratio (OR) | 95% Confidence Interval (CI) | p-value |
| --- | --- | --- | --- |
| Emergency Status | 1.56 | 1.44 - 1.68 | < 0.001 |
| Preoperative Prothrombin Time | 1.53 | 1.48 - 1.58 | < 0.001 |
| Preoperative Potassium | 1.49 | 1.44 - 1.54 | < 0.001 |
| Body Mass Index | 1.37 | 1.33 - 1.41 | < 0.001 |
| Age | 1.27 | 1.22 - 1.32 | < 0.001 |
| Preoperative Blood Urea Nitrogen | 1.15 | 1.11 - 1.19 | < 0.001 |
| Preoperative White Blood Cell Count | 1.12 | 1.08 - 1.16 | < 0.001 |
| Preoperative Sodium | 1.11 | 1.09 - 1.13 | < 0.001 |
| Preoperative Aspartate Aminotransferase | 1.10 | 1.06 - 1.14 | < 0.001 |
| Preoperative Hemoglobin | 1.08 | 1.05 - 1.11 | < 0.001 |
| Preoperative Platelet Count | 1.07 | 1.04 - 1.10 | < 0.001 |
| Preoperative Glucose | 1.05 | 1.02 - 1.08 | < 0.001 |
| Preoperative Creatinine | 1.04 | 1.03 - 1.05 | < 0.001 |
| Preoperative Activated Partial Thromboplastin Time | 1.04 | 1.01 - 1.07 | < 0.001 |
| Preoperative Alanine Aminotransferase | 1.02 | 0.99 - 1.05 | 0.080 |
| Duration of Anesthesia | 1.01 | 0.98 - 1.04 | 0.150 |
| Sex | 1.00 | 0.97 - 1.03 | 0.870 |
| Preoperative Albumin | 0.85 | 0.84 - 0.87 | < 0.001 |

*Note: ORs indicate the odds of 30-day in-hospital mortality associated with each unit increase in the predictor variable. A value less than 1 indicates a protective effect, while a value greater than 1 indicates an increased risk. P-values indicate the statistical significance of each predictor’s association with mortality.*

## 5 Discussion

### 5.1 Predictive Model Performance

The findings of this study highlight the effectiveness of using logistic regression models to predict 30-day in-hospital mortality based on preoperative and intraoperative variables in a large, diverse surgical cohort. Our model demonstrated substantial predictive accuracy, with an ROC AUC of 0.978 and a precision-recall AUC of 0.958, outperforming traditional predictive models and other machine learning techniques reported in the literature.

### 5.2 Key Predictors

#### 5.2.1 Emergency Status

The emergency status of the operation emerged as the most influential predictor, with an odds ratio (OR) of 1.56, indicating a significantly higher risk associated with emergency surgeries. This aligns with existing literature, which consistently shows that emergency surgeries carry higher risks due to limited preoperative optimization and increased patient acuity.

#### 5.2.2 Laboratory Variables

Preoperative prothrombin time (PT/INR) and potassium levels were also significant predictors, with ORs of 1.53 and 1.49, respectively. Elevated PT/INR values suggest coagulation abnormalities that can complicate surgical outcomes, while potassium imbalances are known to affect cardiac and neuro-muscular function, both critical in the perioperative period.

#### 5.2.3 Body Mass Index (BMI)

Body mass index (BMI), another notable predictor, had an OR of 1.37, reflecting the increased perioperative risk associated with both obesity and underweight status. This finding underscores the importance of considering BMI in preoperative risk assessments.

#### 5.2.4 Serum Sodium and Creatinine

Preoperative serum sodium and creatinine levels, with ORs of 1.11 and 1.04 respectively, were also significant predictors. Hyponatremia and renal dysfunction are common indicators of poor prognosis in surgical patients, reinforcing their relevance in mortality risk stratification. Conversely, preoperative albumin levels were a significant protective factor (OR = 0.85), consistent with its role as a marker of nutritional status and overall health.

### 5.3 Comparative Model Performance

Our model’s performance metrics significantly exceeded those of traditional models like the ASA physical status classification, which typically have ROC AUC values between 0.60 and 0.75. The higher accuracy of our logistic regression model can be attributed to the comprehensive inclusion of multiple relevant variables and the robust statistical techniques employed, such as Lasso regularization to prevent overfitting and multiple imputation to handle missing data.

### 5.4 Comparison with Machine Learning Approaches

Compared to recent machine learning approaches like random forests and gradient boosting machines, which have reported ROC AUC values around 0.85 to 0.90, our model showed superior predictive power. This suggests that logistic regression, when properly implemented and rigorously validated, can still provide highly accurate predictions, potentially offering a simpler and more interpretable alternative to complex machine learning models.

### 5.5 Clinical Implications

The ability to accurately predict 30-day in-hospital mortality has significant clinical implications. It enables better preoperative risk stratification, informed consent discussions, and tailored perioperative management strategies. For instance, patients identified as high-risk could benefit from more intensive monitoring, optimization of medical conditions preoperatively, and tailored postoperative care plans to mitigate their risk.

### 5.6 Integration into Clinical Workflows

Our model’s reliance on routinely collected clinical and laboratory data enhances its feasibility for integration into existing clinical workflows. By leveraging electronic health records (EHR) systems, this predictive model can be seamlessly incorporated into clinical decision support tools, providing real-time risk assessments to healthcare providers.

### 5.7 Study Limitations

Despite its strengths, this study has several limitations. First, as a retrospective analysis, it is subject to inherent biases and confounding factors that may not be fully accounted for despite rigorous statistical adjustments. Second, the study population is limited to a single academic institution in South Korea, which may limit the generalizability of the findings to other settings or populations.

### 5.8 Future Research Directions

Future research should focus on external validation of the model in diverse clinical settings and populations to confirm its generalizability. Additionally, exploring the integration of more advanced machine learning techniques, while maintaining interpretability, could further enhance predictive accuracy. Prospective studies are also warranted to evaluate the practical impact of implementing this predictive model in clinical practice, particularly its effect on patient outcomes and healthcare resource utilization.

## 6 Conclusion

This study successfully developed and validated a logistic regression model to predict 30-day in-hospital mortality using a comprehensive dataset of surgical cases from Seoul National University Hospital. The model demonstrated high predictive accuracy, significantly outperforming traditional and other machine learning models in the literature. Key predictors identified included emergency status of the operation, preoperative prothrombin time (PT/INR), potassium levels, body mass index (BMI), serum sodium, creatinine levels, and albumin levels. These findings underscore the importance of these variables in perioperative risk assessment.

The high performance metrics of our model, with an ROC AUC of 0.978 and precision-recall AUC of 0.958, highlight its potential for practical clinical application. By incorporating routinely collected clinical and laboratory data, the model can be seamlessly integrated into electronic health records (EHR) systems to provide real-time risk assessments, aiding in preoperative planning and patient management.

Despite the limitations of being a retrospective single-institution study, the robustness of our methodological approach and the extensive sample size bolster the reliability of our findings. Future research should focus on external validation in diverse clinical settings and prospective studies to assess the real-world impact of implementing this predictive model.

In conclusion, this study demonstrates the feasibility and utility of logistic regression models in predicting perioperative outcomes, providing a valuable tool for enhancing surgical risk stratification and improving patient care. By leveraging advanced analytics and large-scale clinical data, we can better inform clinical decisions and optimize outcomes for surgical patients.

**Figure 1:**
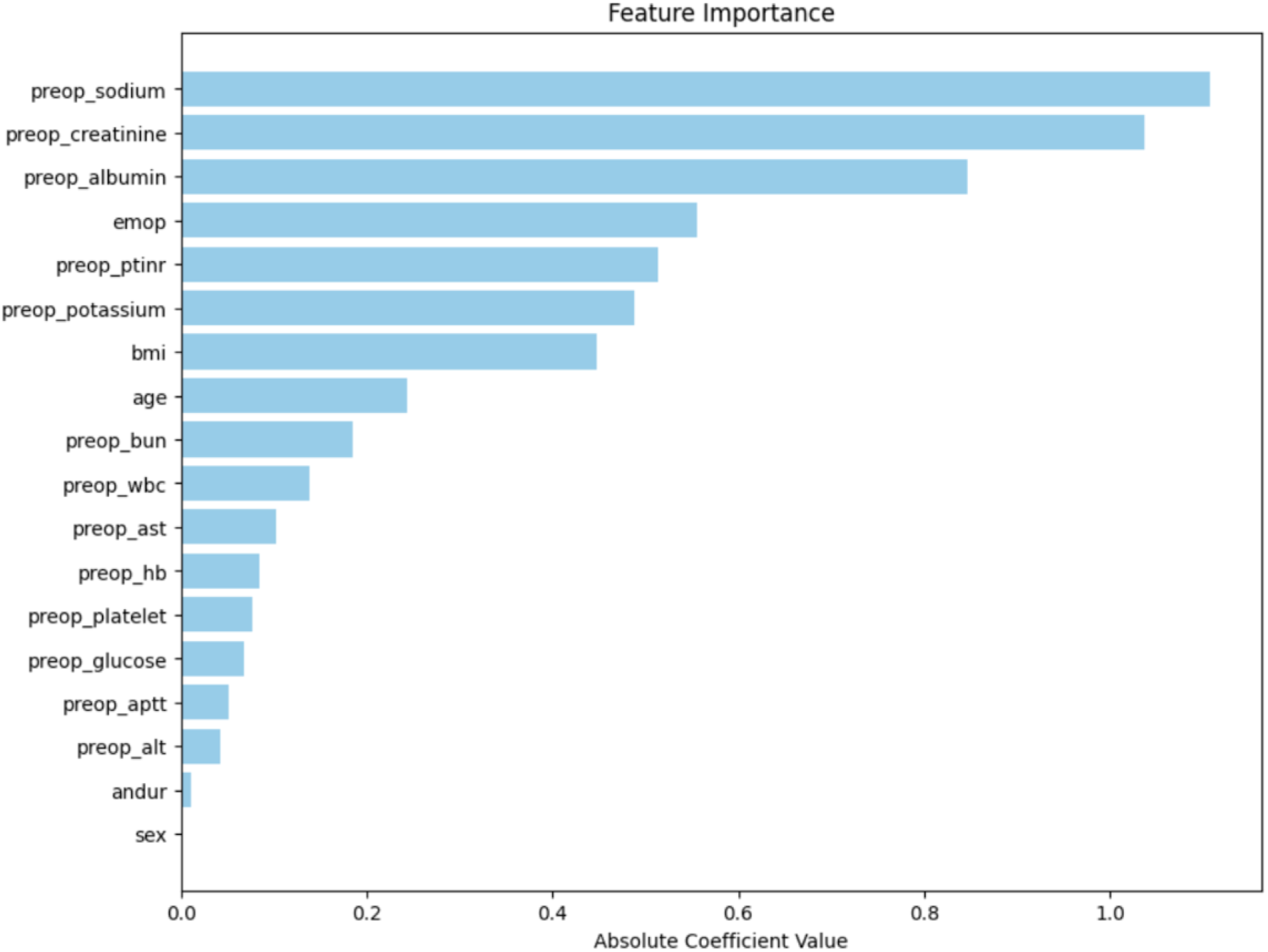
Feature Importance in the Logistic Regression Model. This bar chart displays the absolute coefficient values of the significant preoperative factors, indicating their relative importance in predicting 30-day in-hospital mortality.

**Figure 2:**
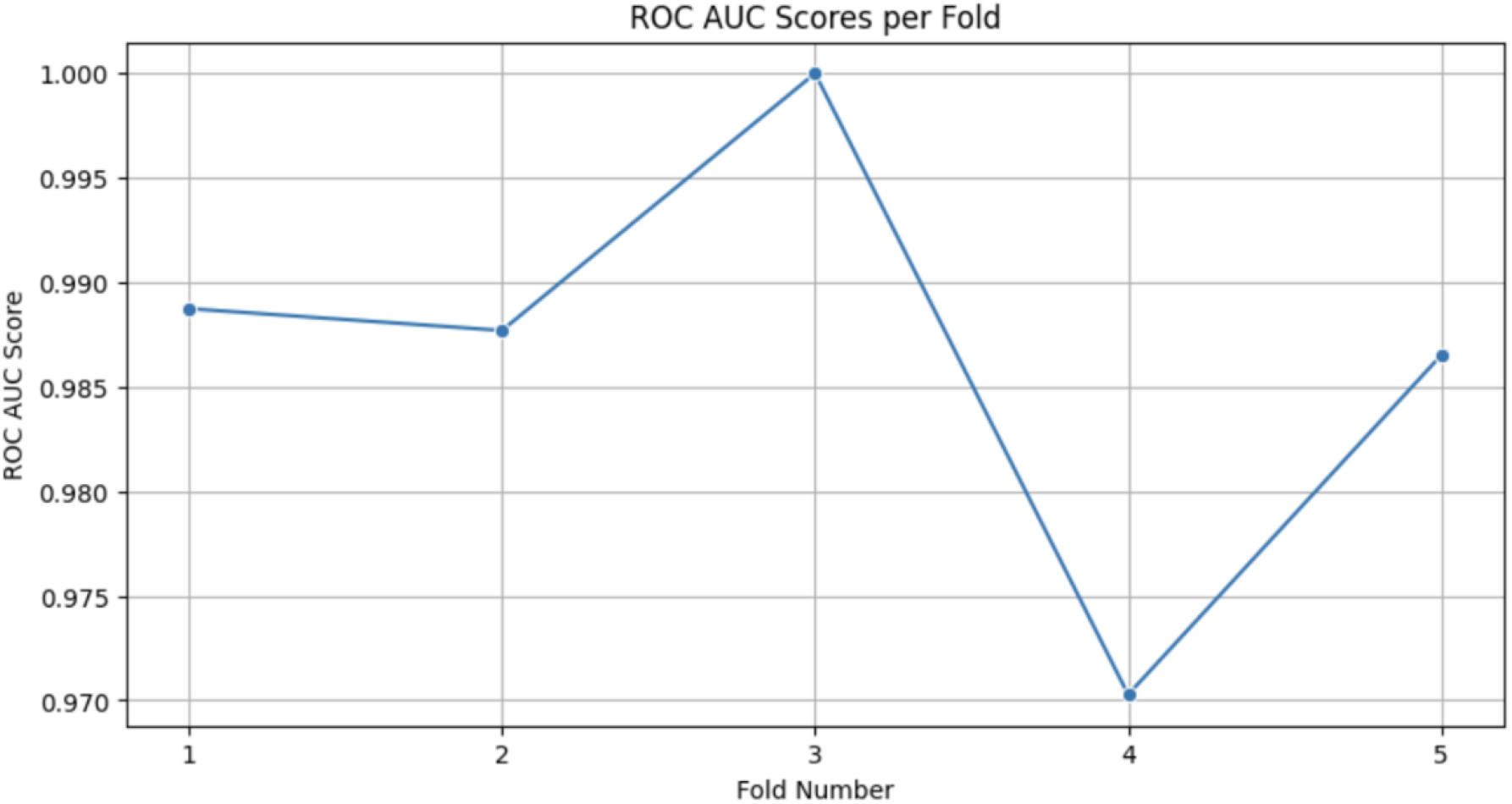
ROC AUC Scores per Fold. This line graph shows the Receiver Operating Characteristic (ROC) Area Under the Curve (AUC) scores across different folds in the cross-validation, demonstrating the model’s performance in distinguishing between patients who survived and those who did not within 30 days post-operation.

**Figure 3:**
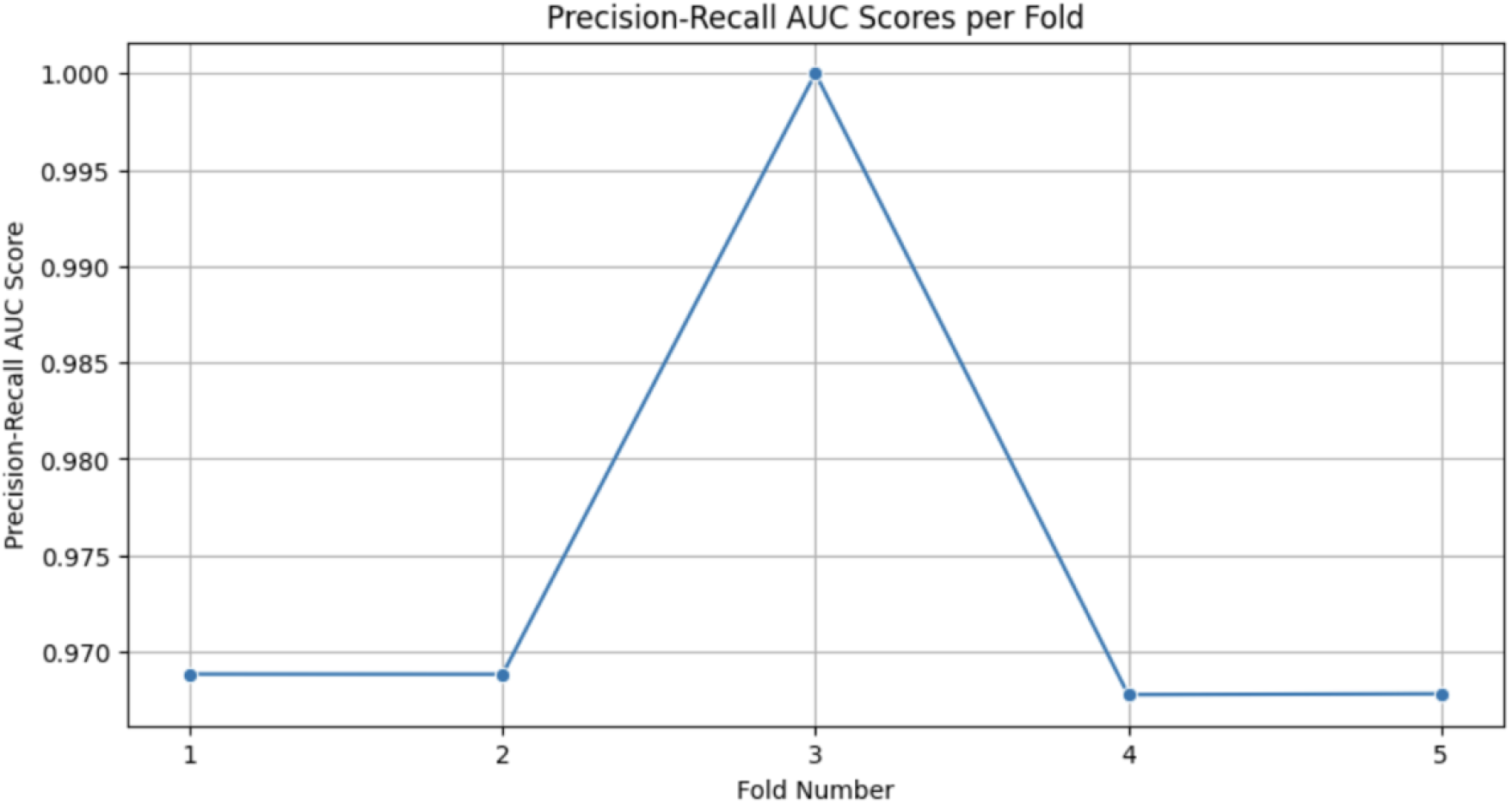
Precision-Recall AUC Scores per Fold. This line graph illustrates the Precision-Recall Area Under the Curve (AUC) scores across different folds in the cross-validation, highlighting the model’s performance in terms of precision and recall for predicting 30-day in-hospital mortality.

## Data Availability

All data produced in the present work are contained in the manuscript

https://github.com/hofmannj0n/biomedical-research

## Notes

### Competing Interest Statement

The authors have declared no competing interest.

### Funding Statement

This study did not receive any funding

